# A deep multimodal model for age-related macular degeneration detection using optical coherence tomography imaging and clinical text data

**DOI:** 10.64898/2026.11.19.26368530

**Authors:** Ming Fang, Hammond Miri, Kerry Bishop, Dorsa Robatjazy, Raymond C. Ko, Ravikrishna Nrusimhadevara, Francisco S. Cayabyab, Seok-Bum Ko, Changiz Taghibiglou

## Abstract

Age-related macular degeneration (AMD) is a major cause of vision loss, with a significant impact on the elderly population worldwide. Machine learning and deep learning methods, combined with optical coherence tomography (OCT), are promising for the rapid diagnosis of AMD. However, most models focus only on one single OCT imaging modality or neglect clinical information. To improve this problem, we design a deep multimodal framework that integrates OCT images with clinical text data for the detection of AMD. Then, we evaluate the proposed model using metrics, such as accuracy, sensitivity, specificity, F-score, and the area under the curve (AUC), achieving excellent detection results at both the image level and patient level. Next, we conduct computational efficiency analysis and independent external validation to further demonstrate the efficiency and generalizability of the proposed model. In addition, we provide visualization results with explainable AI technique to further show the effectiveness of the proposed model. Our proposed model using multimodal strategy involving machine learning along with available clinical text and OCT imaging data accurately predicts individuals with AMD. Therefore, the use of machine learning and explainable AI in AMD detection may ultimately improve early diagnosis and offer more precise and personalized treatment strategies, improving the management of AMD and preserving vision.

## 1. Introduction

Age-related macular degeneration (AMD) is one of the leading causes of visual impairment and blindness, having a great influence on the elderly community in developed countries [1], [2]. AMD is a degenerative condition and affects the macula, which is a central part of the retina [3], [4]. AMD can cause various adverse consequences, such as reading and driving difficulties, as well as a decline in quality of life [5].

The examinations, such as optical coherence tomography (OCT) and OCT angiography (OCTA), play crucial roles in the diagnosis, treatment and management of AMD in clinical practice [6]. OCT and OCTA are non-invasive retinal imaging modalities. OCT provides cross-sectional images of retinal abnormalities, while OCTA captures information on retinal vasculature such as flow density [7]. OCT is the most widely used imaging modality in ophthalmology [8]. OCT images offer rich information on structure and function for retinal diseases such as AMD, which can assist the diagnosis and treatment of such diseases [9].

Previous findings have indicated that the prevalence of AMD shows a growing trend in most countries and regions [10]. In particular, the increase in the elderly population would result in a higher disease prevalence and burden around the world [11], [12]. Some studies have found that about 196 million people globally are suffering from AMD in 2020 [13]. It has been predicted that approximately 288 million people worldwide will be affected by AMD by 2040 [14], [15]. Thus, there is a huge need for early diagnosis and treatment of AMD.

Fortunately, advances in artificial intelligence (AI) and retinal imaging technologies have provided valuable opportunities for the early and rapid detection of retinal diseases, thereby preventing vision deterioration. In recent years, a large number of studies based on machine learning and deep learning have been conducted to predict AMD using OCT and OCTA images.

For example, Rasti et al. [16] proposed an ensemble diagnosis system based on a multi-scale convolutional neural network to classify macular OCT images. The first stage was to adopt a graph-based curvature correction algorithm to remove the distortions in retina and then produce the regions of interest. The second stage was to use a deep ensemble model to recognize normal retina and two common types of macular disease, including AMD and diabetic macular edema. The model evaluated on a public dataset and a private dataset exhibited superior performance in macular OCT classification. Treder et al. [17] applied deep learning to OCT to detect AMD automatically. This work involved a private dataset, containing a total of 1112 OCT images with AMD and a healthy control group. A multi-layer deep convolutional neural network (CNN) pretrained on the ImageNet was trained and tested on the dataset and obtained great performance in AMD identification.

Perdomo et al. [18] designed a deep learning method, called OCT-NET, for classifying retinal diseases including AMD based on OCT data. With a CNN structure, OCT-NET first extracted features from all scans in an OCT volume to identify each B-scan as AMD or healthy. Then, the proposed method was evaluated on the public A2A OCT dataset from the Age-Related Eye Disease Study 2 (AREDS2), attaining high values in terms of sensitivity, specificity, accuracy and the area under curve (AUC). Rong et al. [19] developed a surrogate-assisted model for the automatic recognition of retinal OCT images. Specifically, noise in the input OCT images was reduced using the denoising technique. Thresholding and morphological dilation were then used to obtain the masks. After that, the denoised images and the masks were utilized to produce the surrogate images. Next, these surrogate images were sent to a CNN for training. Given an input image, its final prediction result was decided by the average of the outputs from the CNN model based on the surrogate images. The proposed model was tested to distinguish between AMD, diabetic macular edema and healthy controls, achieving an accuracy of 0.9509 and an AUC of 0.9856 on the public Duke dataset.

Yim et al. [20] introduced a deep learning network to predict progression to AMD based on OCT images and automatic tissue maps. The experimental results showed that the proposed method obtained good sensitivity and specificity, which illustrated the feasibility of applying artificial intelligence techniques to disease progression prediction. Romo-Bucheli et al. [21] built a deep learning model to predict treatment requirements for AMD from retinal OCT images. Specifically, the model consists of a densely connected neural network (DenseNet) and a recurrent neural network (RNN), in which DenseNet learns retinal spatial features and RNN combines information from different time points. The proposed model performed better than existing machine learning methods, indicating its ability to extract effective patterns from OCT images.

Recently, Zang et al. [22] established an automated diagnostic framework based on semi-sequential 3-dimensional (3D) CNN using structural OCT and OCTA data volumes. In this framework, 526 OCT and OCTA volumes were scanned from the eyes of 91 healthy participants, 95 patients with AMD, 161 patients with diabetic retinopathy, and 108 patients with glaucoma. First, the 3D OCT and OCTA scans were combined and then were inputted to a semi-sequential classifier. Finally, the inputs were classified as healthy, AMD, diabetic retinopathy, or glaucoma. The framework yielded reliable results in retinal disease detection, especially with an AUC of 0.98 for the diagnosis of AMD.

Bui et al. [23] used a multi-scale learning method and a sparse residual network to diagnose multiple retinal diseases such as AMD from OCT images. Their multi-scale method captured features from different sizes of OCT images, while the sparse residual network with smaller convolution size decreased model complexity. The proposed method performed well, showing an accuracy of 0.9740, a sensitivity of 0.9538, and a specificity of 0.9825 on a private dataset. Zhou et al. [24] proposed a foundation model, named as RETFound, for generalizable disease detection from retinal images. First, RETFound was trained on a large number of unlabelled retinal images and then adapted to disease detection. Moreover, RETFound was validated on several datasets, including OCT data for multiple disease classification and prognosis such as AMD, yielding great performance compared with the competing methods.

Xu et al. [6] constructed a bi-modal deep learning structure for the detection of AMD and polypoidal choroidal vasculopathy. Specifically, both OCT and color fundus images were inputted into two deep CNN models to extract features, respectively. Then, the two features were combined, and specific categories were generated in the end. The proposed bi-modal structure showed excellent results, with an accuracy of 0.874, an F1-score of 0.878, a sensitivity of 0.888, and a specificity of 0.956 on a private dataset. Similarly, Wang et al. [25] developed a two-stream CNN model to combine OCT and color fundus photograph, obtaining great classification performance for AMD.

These models have reached good performance and can serve as helpful tools for detecting AMD. However, most of studies only apply a single OCT imaging modality to deep learning models. Although some studies have taken multiple modalities as the input, such as the one by Xu et al. [6], they have not taken age information into consideration. Furthermore, previous research has shown that the development of AMD is closely related to ageing [14], [10].

Therefore, we develop a deep multimodal integration model to fuse OCT images with clinical text information involving patient ages for a complete understanding of the disease. Then, we apply the proposed model on both public and private OCT datasets. The experimental results show that the proposed model achieves excellent performance both at the image level and at the patient level in accuracy, precision, recall or sensitivity, F-score, specificity, and AUC values. In addition, we illustrate the model performance with ROC curves, confusion matrices, and an explainable AI technique. These visualization results demonstrate the detection ability of the proposed model for the disease. We conduct computational efficiency analysis and external validation to further illustrate the efficiency and generalizability of our model. The proposed model is expected to provide effective screening and diagnosis of AMD at an early stage, thereby assisting in preventing poor visual outcomes.

## 2. Datasets

### 2.1. Public Dataset

The OCT dataset for image-based deep learning (OCTDL) [26] was applied to develop the deep multimodal integration model. Some sample images from the OCTDL are displayed in Figure 1. The OCTDL dataset consists of 821 subjects, including clinical text information, such as patient ID, disease labels, sex and year of birth, and OCT images with seven classes, as shown in Table 1.

**Table 1.** The details of OCTDL dataset.

| Disease | Label | Number of Scans | Number of Patients |
|---|---|---|---|
| Age-related Macular Degeneration | AMD | 1231 | 421 |
| Diabetic Macular Edema | DME | 147 | 107 |
| Epiretinal Membrane | ERM | 155 | 71 |
| Normal | NO | 332 | 110 |
| Retinal Artery Occlusion | RAO | 22 | 11 |
| Retinal Vein Occlusion | RVO | 101 | 50 |
| Vitreomacular Interface Disease | VID | 76 | 51 |
| Total |  | 2064 | 821 |

**Figure 1.**
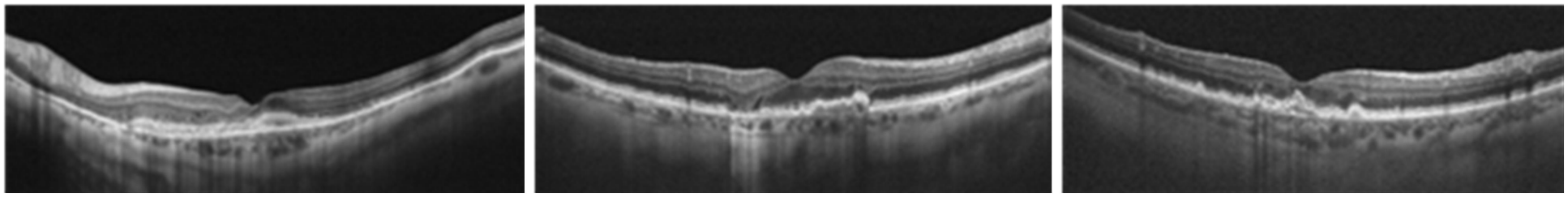
Sample images from OCTDL dataset

This study mainly focuses on detecting class AMD and therefore formulates the problem as binary classification ask, that is, AMD and NON-AMD.

### 2.2. Independently Collected Clinical Dataset

To assess the generalization capability of the proposed model beyond the development dataset, we collected an independent external cohort from local clinical practice settings, including clinics in Saskatoon and Regina. This cohort comprises 40 subjects, including 20 patients with AMD and 20 age- and sex-matched healthy controls. All available images and corresponding clinical metadata were retained for evaluation. Crucially, none of this data was used in model training, development, or internal validation. This external cohort was specifically utilized to assess the performance of our model on data obtained in an independent, real-world clinical environment.

## 3. Methods

In this section, data preprocessing and the proposed model are described in detail. The overall workflow of the model is depicted in Figure 2. The first stage was to input OCT images and clinical text data into an Image Encoder and a Text Encoder, respectively, to learn feature representations. The second stage was to develop a Merge Module to fuse these features for disease prediction.

**Figure 2.**
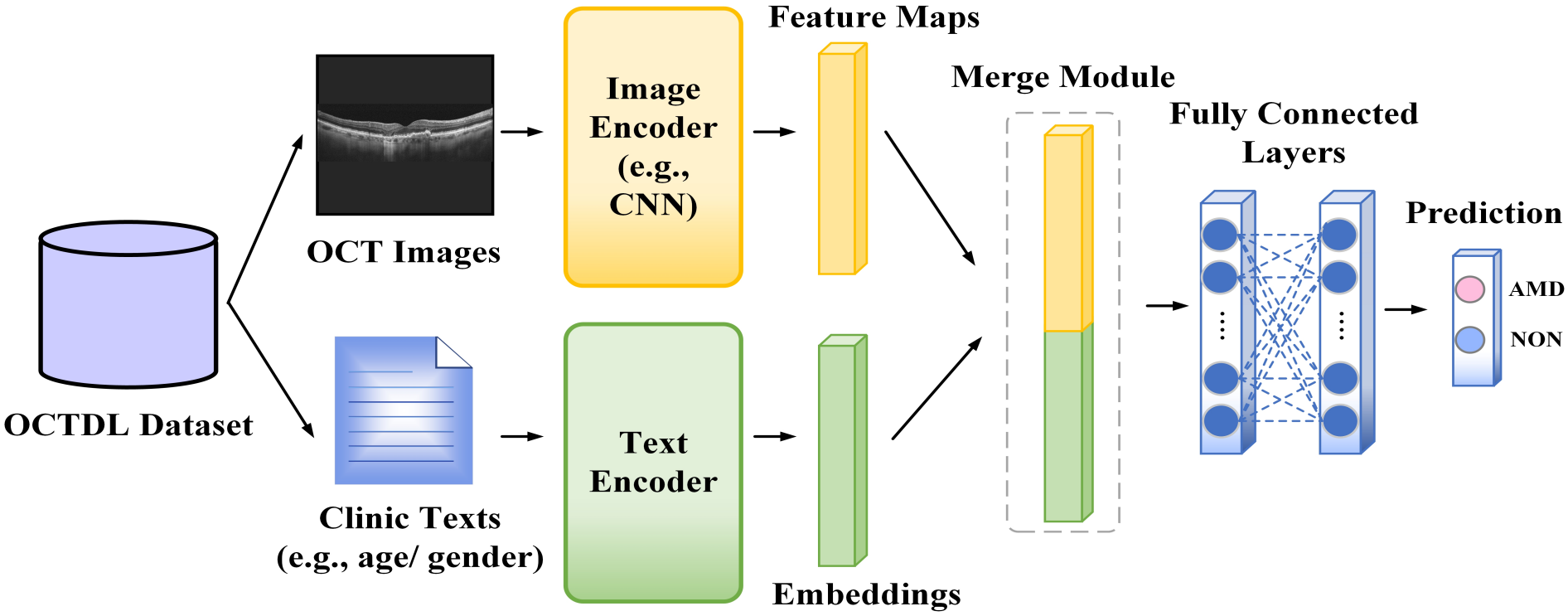
The workflow of the deep multimodal detection model

### 3.1. Data Preprocessing

#### 3.1.1. Age and Sex Preprocessing

The dataset contains invalid values for age and sex. Therefore, the missing age and sex were filled with reasonable values. Specifically, missing age values were imputed using the median age calculated from the training set. If the training set contained no valid age values, a predefined value of 50 was used. An indicator for missing age was retained to explicitly inform the model whether the original age value was unavailable. Subsequently, the imputed age was standardized using z-score normalization to stabilize model training. Furthermore, unavailable or unrecognized sex values were treated as missing. Sex was processed using one-hot encoding, with missing sex represented as a separate category. A sex missing indicator was also retained in the one-hot representation.

#### 3.1.2. Five-fold Cross Validation & Patient-level Split

The patients were assigned to folds and the number of classes was balanced in each fold. Specifically, the data were grouped by patient ID, and then these patient groups were split into several folds.

#### 3.1.3. Imbalanced Dataset

There are fewer samples in some classes, which may lead to bias. In order to address this issue, a weighted sampling approach was implemented during model training to ensure balanced sampling. Specifically, each sample was assigned a weight inversely proportional to the frequency of its majority class. This approach increased the sampling probability of minority classes, thus mitigating class imbalance. As a result, samples from underrepresented classes were sampled more frequently during training, reducing the bias toward dominant classes.

### 3.2. Model Design

The features of clinical text data are important biomarkers in OCT images for the prediction of AMD. In order to combine OCT images with clinical texts, a deep multimodal integration model was designed.

There are four main components in the proposed deep multimodal model (Figure 2): an image encoder, a text encoder, a merge module, and a prediction module, which are described in detail below.

#### 3.2.1. Image Encoder

Convolutional neural network (CNN) models are a significant part in deep learning. In general, a CNN consists of convolutional layers, pooling layers, and fully connected layers. The main purpose of convolutional and pooling layers is to extract features from the input data. Fully connected layers are used for classification and prediction based on the extracted features.

CNN models have been successfully applied in a wide range of medical problems [27], [28], [29] and are seen as important tools to help clinicians and neurologists predict diseases. One of the CNN models, ResNet [30], has special residual blocks that can greatly enhance model performance. Here, ResNet18 was employed as the image encoder to extract feature maps from the input OCT images.

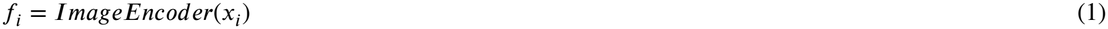

where *f*_*i*_ represents the features obtained by the image encoder from OCT images *x*_*i*_.

ResNet18 was chosen because it provided a good balance between predictive performance and computational cost compared with deeper or larger CNN architectures. The selected ResNet18 backbone was truncated before its original classification layer. Average pooling was applied to the resulting convolutional feature map, followed by a fully connected projection layer that mapped a 512-dimensional representation to a 256-dimensional image embedding. This embedding was subsequently fused with the clinical metadata representation.

#### 3.2.2. Text Encoder

Multilayer perceptron (MLP) is a machine learning algorithm consisting of an input layer, one or multiple hidden layers, and an output layer [31], [32]. MLP algorithms have proven to be effective in text analysis [33], [34], [35]. Here, the MLP was adopted as the text encoder to learn textual embeddings from input clinical information, including age and sex of patients.

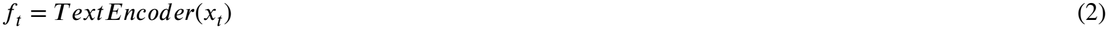

where *f*_*t*_ denotes the embedding vectors obtained by the text encoder from the clinical information *x*_*t*_.

The input consisted of five features: standardized age, an age missing indicator, sex indicators for male or female, and missing sex. The resulting five-dimensional vector was processed by a fully connected layer with 128 hidden units, followed by a ReLU activation. A final fully connected layer projected the representation to a 64-dimensional clinical feature embedding. The encoded clinical representation was subsequently integrated with the 256-dimensional image representation.

#### 3.2.3. Merge Module

After capturing features from OCT images and clinical text data with the image encoder and text encoder, respectively, a merge module was constructed through concatenation. The concatenation methods can fuse information from multiple data sources, thereby generating more diverse and effective representations for disease prediction, which have received attention in various applications [36], [37], [38].

The merge module can combine features from the two modalities for richer representations. Specifically, the encoded clinical representation was integrated with the image representation. The merged features *f*_*m*_ can be defined as follows.

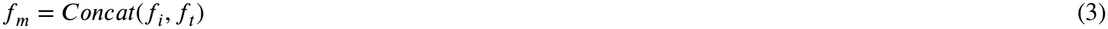

where Concat(·) represents the concatenation operation, aiming to concatenate two types of features into one single feature.

#### 3.2.4. Prediction Module

The merged features were fed into the prediction module for final disease detection. The prediction module is composed of fully connected layers and a prediction head. Specifically, a *c*-dimensional vector was outputted after passing fully connected layers, where *c* means the number of classes. Therefore, specific categories of the disease can be produced.

Finally, the proposed deep multimodal method can determine whether the input is AMD or NON-AMD.

#### 3.3. Training Details

In this study, the Adam, an adaptive optimization algorithm, was set as the optimizer to train the proposed model. The model was trained 100 epochs with a learning rate of 0.0001 and a batch size of 16 on an NVIDIA A100 GPU. Moreover, five-fold cross validation was adopted. In this strategy, the dataset was divided into five parts. Four parts were used for training, and the remaining part was used for testing. The model was trained and evaluated on the training and testing sets, respectively. This process was repeated five times, and the average of the five results was taken as the final performance evaluation for the model.

## 4. Experimental Results

In this section, the evaluation metrics used in this study and the experimental results are reported.

### 4.1. Evaluation Metrics

In disease prediction tasks, widely used evaluation metrics contain accuracy, precision, recall or sensitivity, F-score, and specificity. Accuracy is represented as the proportion of correctly predicted samples out of the total sample size. Precision indicates the proportion of true positive samples out of the total number of positive predictions. Recall or sensitivity expresses the proportion of true positive samples out of all positive samples. F-score is denoted as the harmonic mean of precision and recall. Specificity refers to the proportion of true negative samples out of all negative samples. The specific formulas are written as follows.

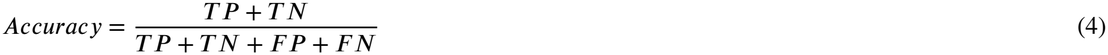

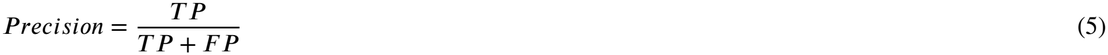

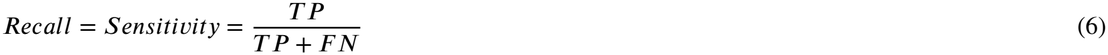

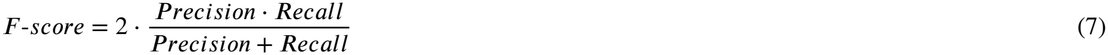

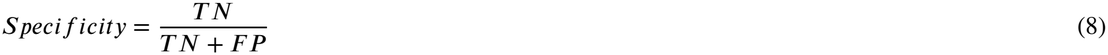

where TP (true positive) is the number of positive samples that are correctly predicted. FP (false positive) is the number of negative samples that are incorrectly recognized as positive. TN (True negative) is the number of negative samples that are correctly detected. FN (false negative) is the number of positive samples that are mistakenly predicted as negative.

In addition, the model performance was demonstrated through the Area Under the Receiver Operating Characteristic Curve (AUC) with 95% confidence intervals (CIs), the Receiver Operating Characteristic (ROC) curve, confusion matrix, and gradient-weighted class activation mapping (Grad-CAM).

### 4.2. Results and Discussion

Based on the combination of OCT images and clinical metadata, the proposed model can capture better feature representations for disease prediction. We display both image-level and patient-level detection results. From a series of experiments, we observed that the proposed model can yield great results on the OCTDL dataset at the image level and at the patient level. Further evaluation and analysis were conducted subsequently.

#### 4.2.1. Image-level

We assessed the performance of the model on the OCTDL dataset through quantitative analysis, including accuracy, precision, recall or sensitivity, F-score, and specificity. As shown in Figure 3, the model achieved high values in terms of all metrics at the image level. In particular, the accuracy and F-score values of the proposed method were both above 0.952. Notably, both the sensitivity and specificity of the model were close to 0.956, which is especially important for identifying positive cases in clinical tasks.

**Figure 3.**
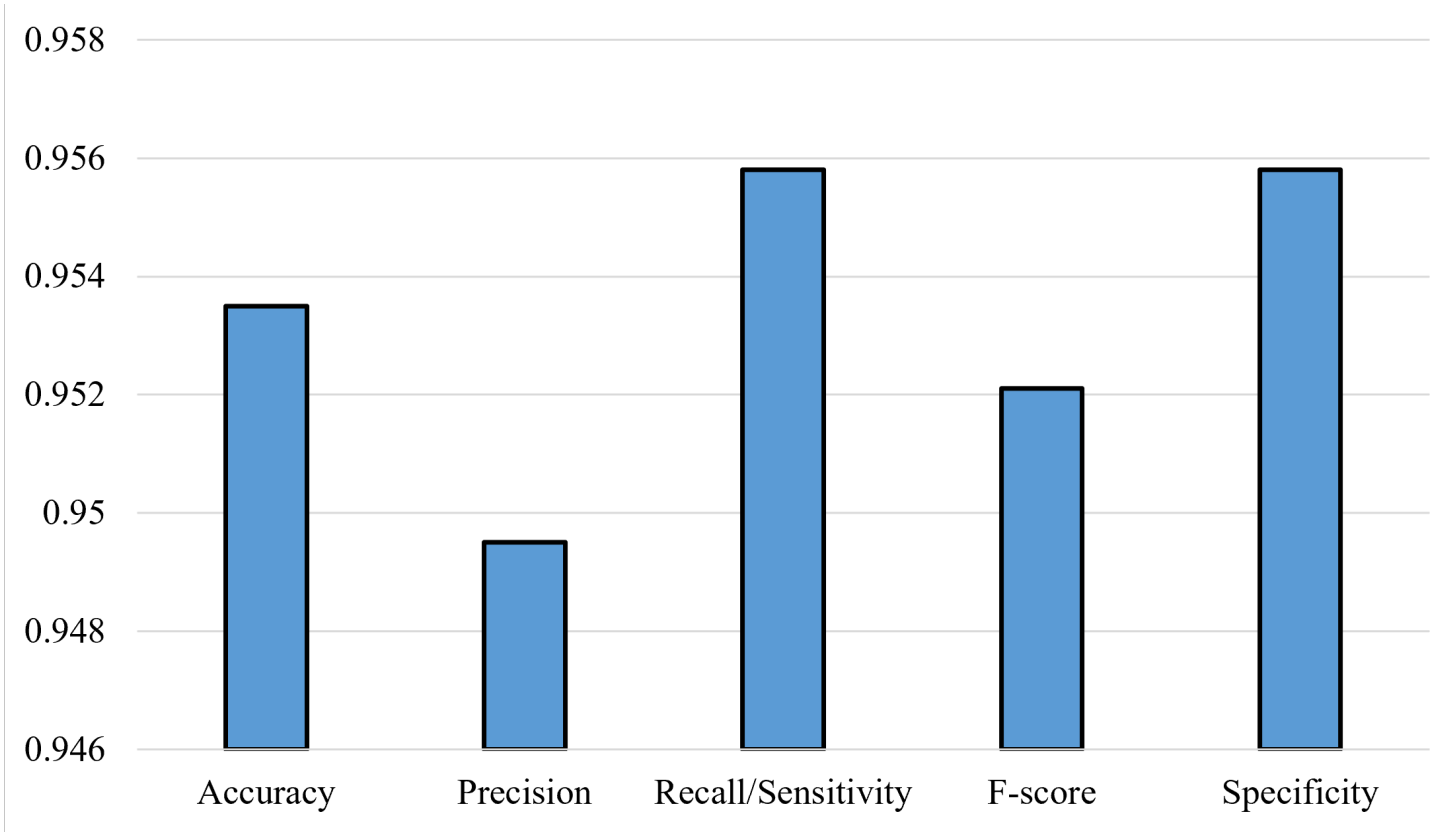
Quantitative results at the image level on OCTDL dataset

AUC and ROC are generally adopted to measure the discriminative ability of models for predictive purposes [39], [40]. Models with higher AUC values are beneficial for clinical applications. In other words, higher AUC values refer to better discrimination between high-risk and low-risk patients [39]. The ROC curve can effectively visualize the performance of a model [41]. From the ROC curve in Figure 4, the AUC value with 95% CI was 0.9854 (0.9808-0.9894), which indicated that the model obtained great and stable image-level performance.

**Figure 4.**
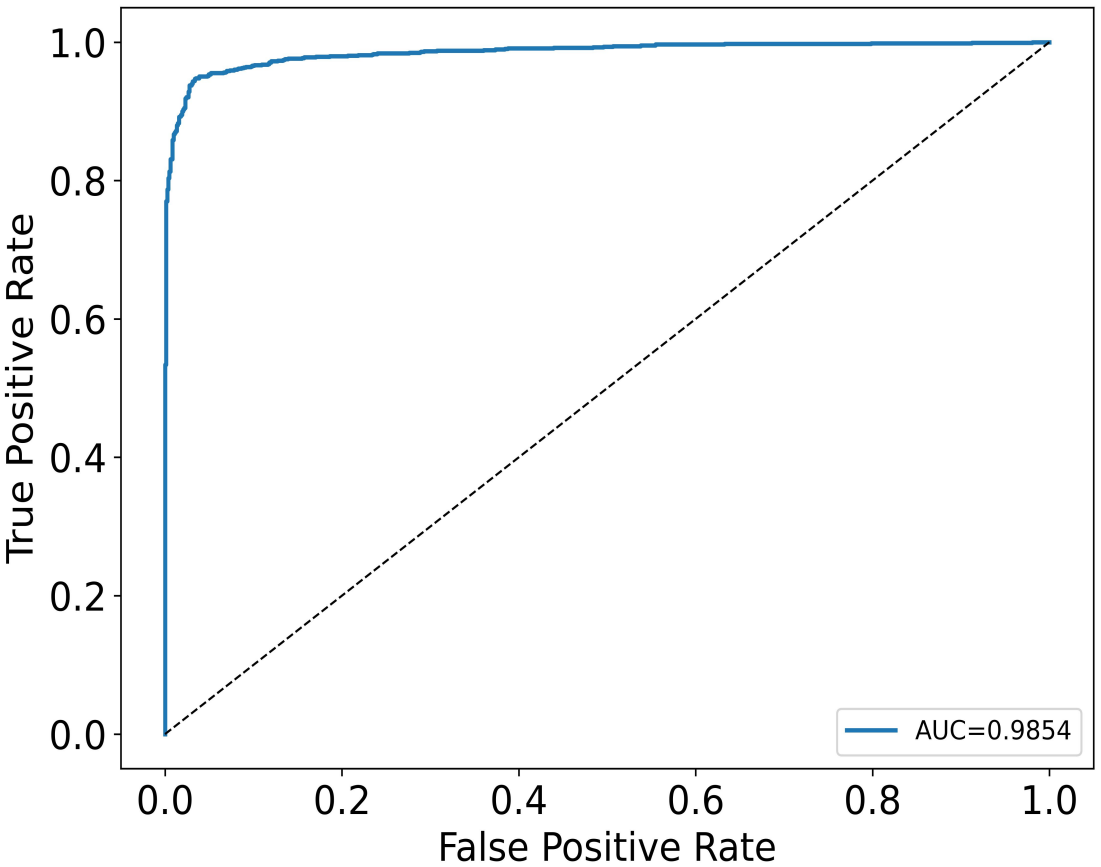
ROC curve at the image level on OCTDL dataset

As demonstrated in the confusion matrix (Figure 5), the proposed model was able to correctly predict the majority of samples. Specifically, the model accurately identified 1162 images out of a total of 1231 images for AMD samples and 806 images out of 833 images for NON-AMD samples.

**Figure 5.**
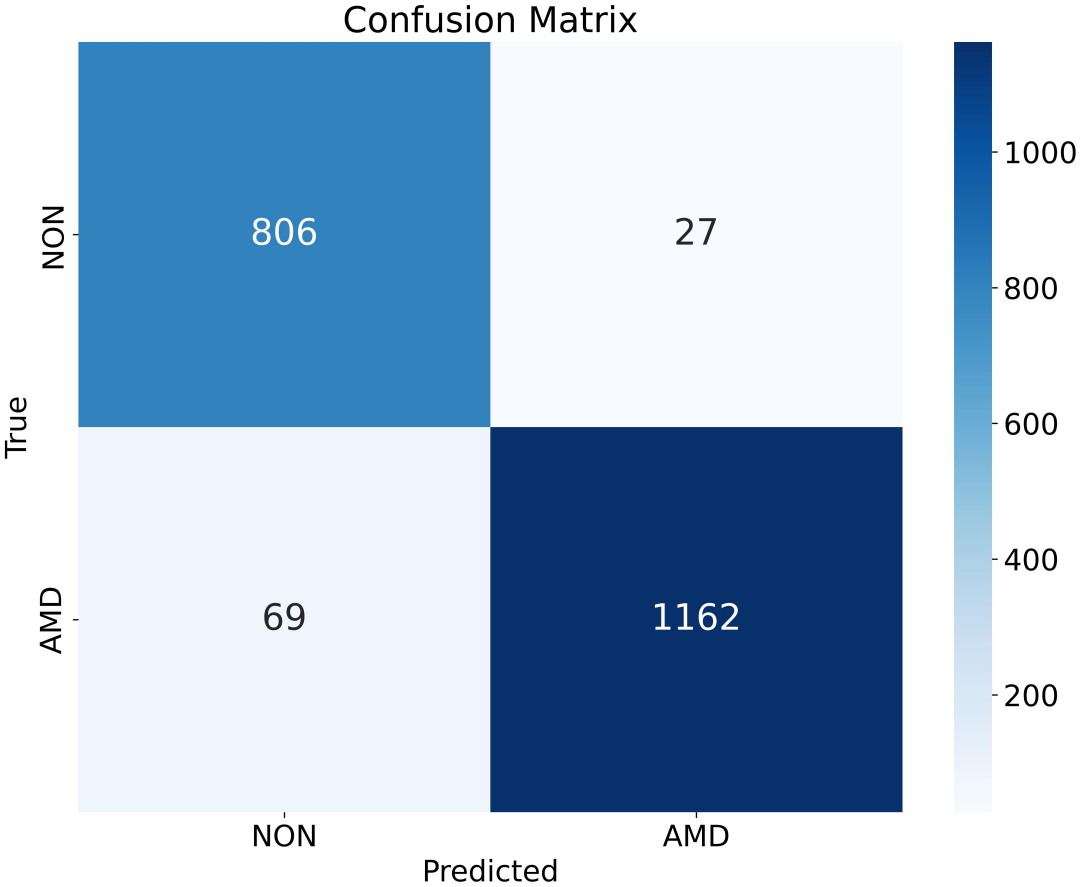
Confusion matrix at the image level on OCTDL dataset

#### 4.2.2. Patient-level

Patient-level results are more meaningful in real-world clinical applications. To obtain results at the patient level, the predictions of all images that belong to the same patient are aggregated into a prediction at the patient level by majority voting, a scheme for decision making [42]. In majority voting, the final prediction of a patient is decided by the most common prediction among all image-level predictions.

Similarly, we also report the quantitative results at the patient level. As shown in Figure 6, the proposed model performed excellently at the patient level. We found that our proposed model surpassed approximately 0.942 in terms of accuracy and F-score. The model also achieved great values of precision, sensitivity, and specificity, with values exceeding 0.943.

**Figure 6.**
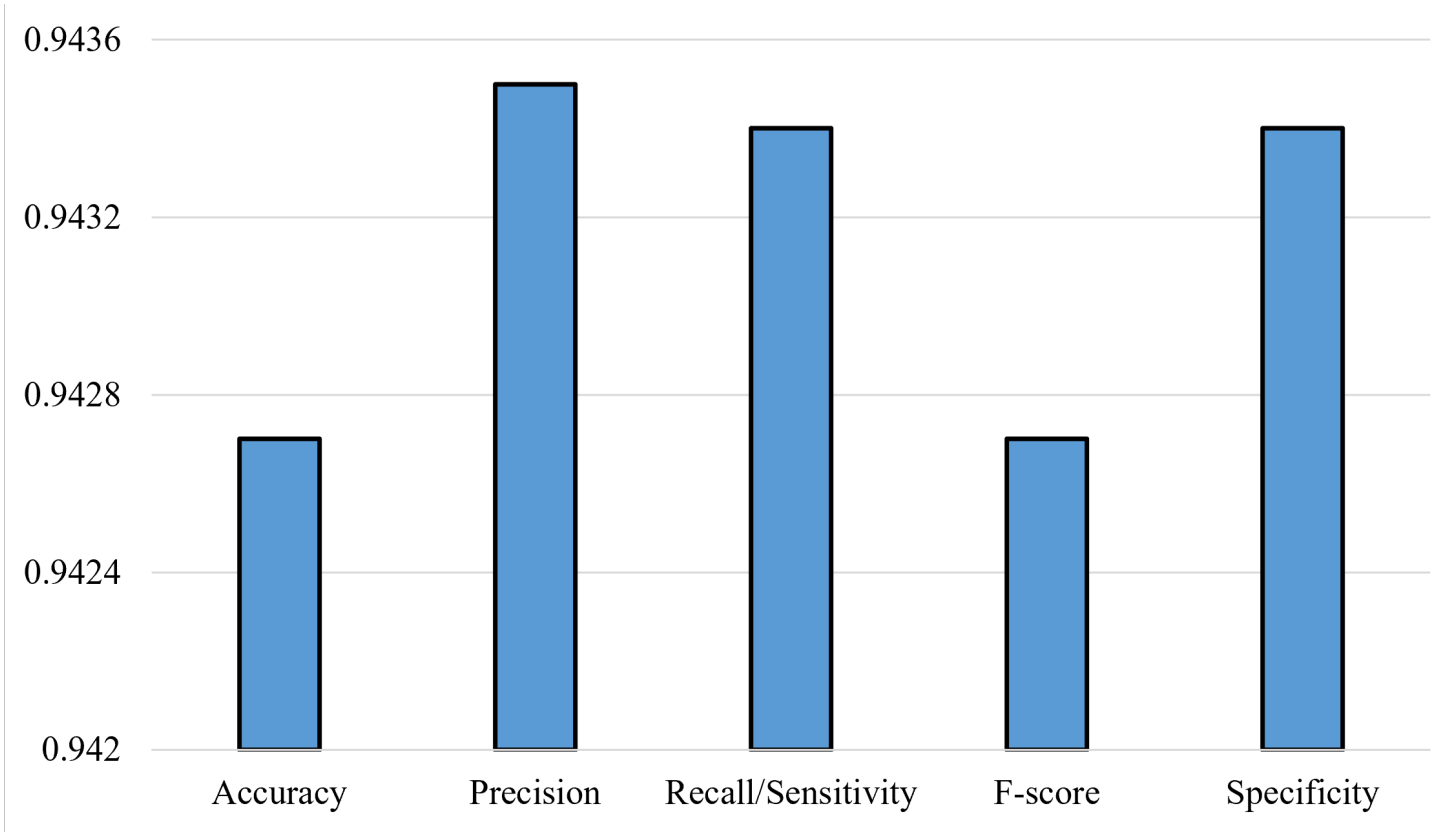
Quantitative results at the patient level on OCTDL dataset

From the ROC curve in Figure 7, the AUC with 95% CI was 0.9855 (0.9777-0.9920), which demonstrated the effectiveness of the model at the patient level.

**Figure 7.**
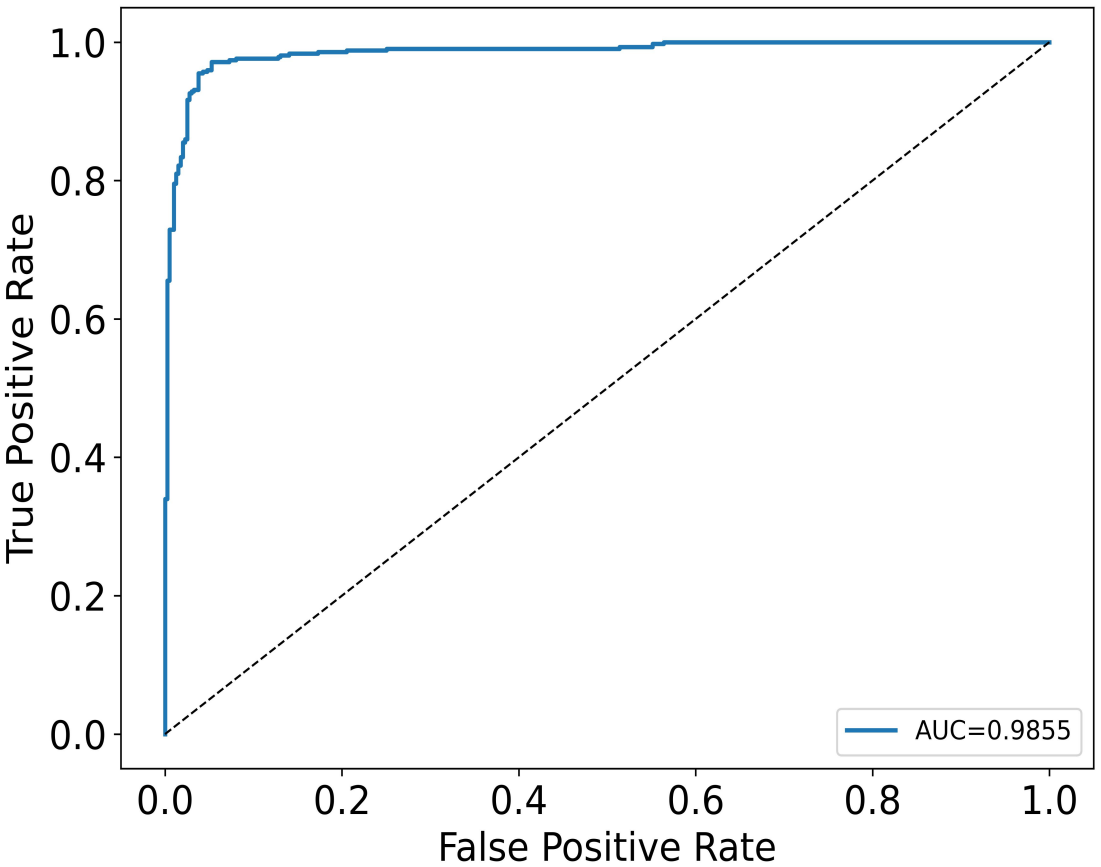
ROC curve at the patient level on OCTDL dataset

As shown in the confusion matrix (Figure 8), most of the patients were correctly predicted by the proposed model. Specifically, the proposed model correctly detected 387 individuals with AMD and 386 individuals with NON-AMD.

**Figure 8.**
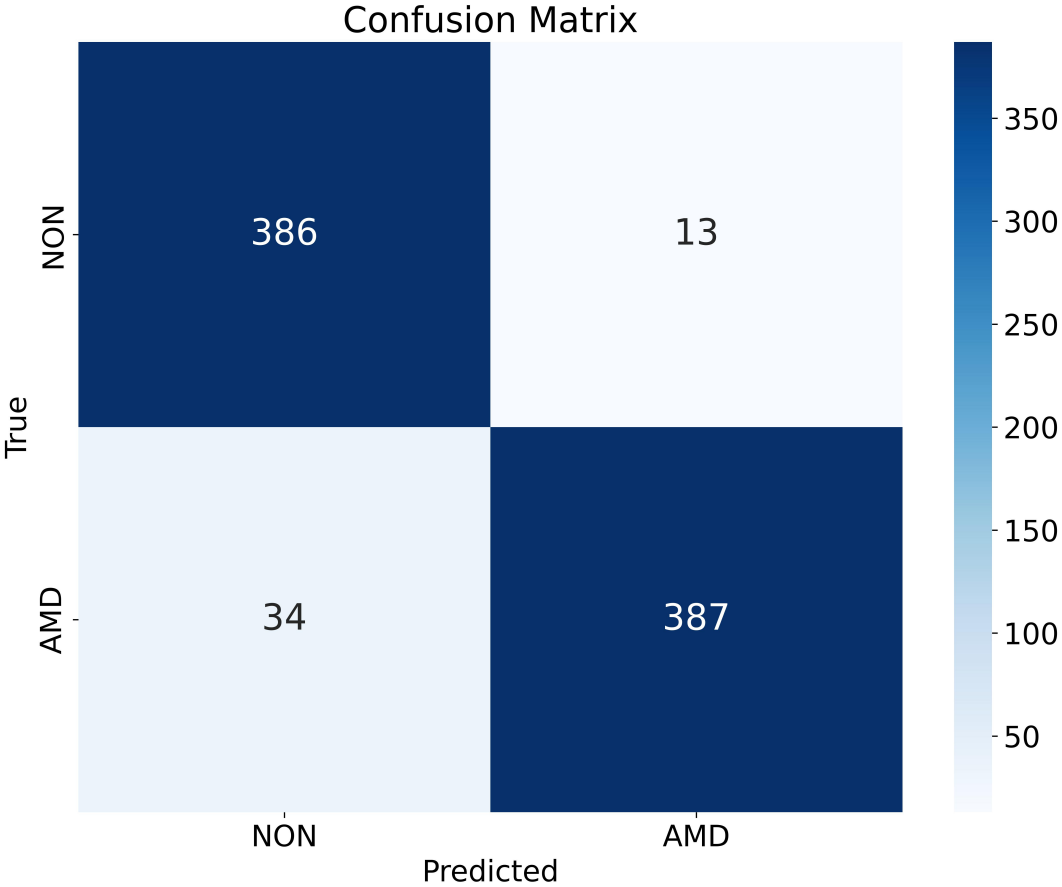
Confusion matrix at the patient level on OCTDL dataset

#### 4.2.3. Ablation Analysis

To validate the effectiveness of our multimodal strategy, we compared the proposed method (image + text) with image only model and text only model at both the image level and patient level. All models (image only, text only, and multimodal) were evaluated using the same patient-level aggregation approach, cross-validation splitting, evaluation metrics, and statistical procedures. The results, including accuracy (ACC), precision (PRE), recall (REC) or sensitivity (SEN), F-score (F), specificity (SPE), and AUC, are shown in Table 2 and Table 3. All highest values are displayed in bold for clarity.

**Table 2.** Ablation analysis at the image level on OCTDL dataset.

| Model | Image | Text | ACC | PRE | REC/SEN | F | SPE | AUC |
|---|---|---|---|---|---|---|---|---|
| Image Only | ✓ | × | 0.9302 | 0.9282 | 0.9338 | 0.9281 | 0.9338 | 0.9788 |
| Text Only | × | ✓ | 0.9307 | 0.9284 | 0.9346 | 0.9287 | 0.9346 | 0.9778 |
| **Proposed method** | ✓ | ✓ | **0.9535** | **0.9495** | **0.9558** | **0.9521** | **0.9558** | **0.9854** |

**Table 3.**
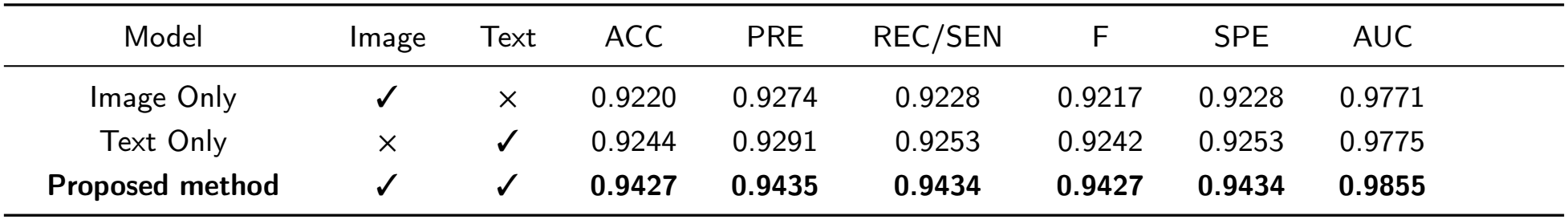
Ablation analysis at the patient level on OCTDL dataset.

As shown in Table 2, the contribution of clinical metadata was evaluated through an image only versus multimodal ablation using the same ResNet18 backbone. The image only model achieved an accuracy of 0.9302, a precision of 0.9282, a recall of 0.9338 and an F-score of 0.9281 at the image level. Compared with the image only model, our proposed model had improvements of 0.0233, 0.0213, 0.022, and 0.024 in accuracy, precision, recall, and F-score, respectively. Compared with the text only model, our proposed model increased by 0.0228, 0.0211, 0.0212, and 0.0234 in terms of accuracy, precision, recall, and F-score, respectively.

As shown in Table 3, all metrics results of our proposed method were more than 0.94 at the patient level. However, compared with the proposed model, the image only model decreased by approximately 0.021, 0.016, 0.021, and 0.021 in accuracy, precision, recall, and F-score, respectively. Similarly, our proposed multimodal model performed better than the text only model, which improved performance by approximately 0.018 and 0.019 in sensitivity and F-score, respectively. From the above ablation analysis, the proposed method attained great performance in all metrics due to the adoption of a multimodal strategy. It is clear that the proposed multimodal method is more effective than the single modality in AMD prediction. Our method benefits from both imaging and clinical metadata, in which the former provides substantial discriminative information, while the latter contributes valuable predictive information.

#### 4.2.4. Comparison with Other Methods

The method proposed in this study focused specifically on identifying AMD category. To further demonstrate the detection capability of our proposed method for AMD, we compared it with some existing methods recently published on the OCTDL dataset for predicting AMD category, including those proposed by Aouti et al. [43], Kim et al. [44], and Naim et al. [45]. Here, the performance regarding the target category, AMD, was examined. Specifically, ACC, PRE, REC/SEN, F, and SPE were compared when these metrics were available from these competing studies. The comparison results are presented in Table 4, with the highest values indicated in bold.

**Table 4.**
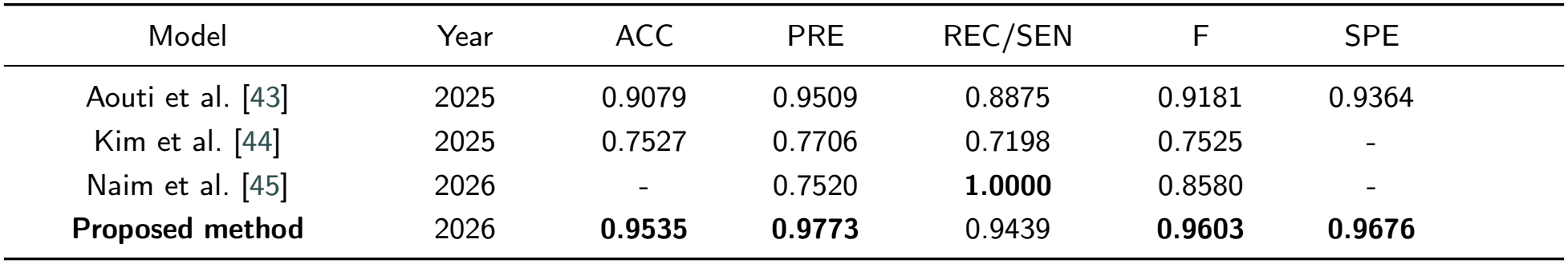
The comparison results on OCTDL dataset for AMD detection.

As reported in Table 4, Aouti et al. [43] proposed a lightweight multi-scale attention model for classifying OCT images, which acquired a precision of 0.9509 and an F-score of 0.9181. The method proposed by Aouti et al. [43] had the second best performance, but its sensitivity value was lower than those of the proposed method and the method of Naim et al. [45]. Specifically, our proposed method outperformed the method of Aouti et al. [43], with improvements of approximately 0.05, 0.03, 0.06, 0.04, and 0.03 in accuracy, precision, sensitivity, F-score, and specificity, respectively. Although Kim et al. [44] introduced a modified variational autoencoder model for AMD prediction, all evaluation metrics of this model were significantly smaller than those of the proposed method. Compared to the method of Kim et al. [44], our proposed method had an increase of roughly 0.20, 0.21, 0.22, and 0.21 in terms of accuracy, precision, sensitivity, and F-score, respectively. In the work from Naim et al. [45], a capsule network framework was presented for retinal disease classification. The work of Naim et al. [45] yielded the highest sensitivity value among all methods, yet its precision and F-score were much lower than those of the proposed method. Specifically, our proposed method rose precision by 0.23 and F-score by 0.10 compared to the work of Naim et al. [45].

According to a series of experimental analysis, we observed that most existing methods obtained the prediction results of AMD using one single modality. Benefitting from a multimodal strategy, our proposed model can achieve outstanding performance among all competing methods for the identification of AMD disease on the OCTDL dataset, thereby obviously demonstrating the superiority of our proposed multimodal method in AMD detection.

#### 4.2.5. Computational Efficiency Analysis

To assess the computational efficiency of the proposed method, we compared the number of network parameters and floating-point operations (FLOPs) across the proposed method and competing models, including ResNet34, ResNet50, VGG16, VGG19, EfficientNet-B0, and MobileNetV3. All models were trained and evaluated on the same OCTDL dataset as the proposed method.

As shown in Table 5, the proposed method contains approximately 11.36M parameters and requires approximately 18.98 GFLOPs, with the highest accuracy, precision, F-score, sensitivity, and specificity compared with other competing models. The proposed method uses fewer parameters and requires fewer computational operations than ResNet34, ResNet50, VGG16, and VGG19. In particular, it reduces the parameter count by approximately 9.93M, 12.15M, 122.91M, and 128.22M, relative to ResNet34, ResNet50, VGG16, and VGG19, while maintaining comparable performance. Although the proposed method employs a little more parameters than EfficientNet-B0 and MobileNetV3, it achieves more superior performance. These results indicate that the proposed architecture achieves a favorable balance between model complexity and predictive capability.

**Table 5.** Comparison results of different methods on OCTDL dataset.

| Model | Params(M) | FLOPs(G) | ACC | PRE | REC/SEN | F | SPE | AUC |
|---|---|---|---|---|---|---|---|---|
| ResNet18 | 11.18 | 18.98 | 0.9302 | 0.9282 | 0.9338 | 0.9281 | 0.9338 | 0.9788 |
| ResNet34 | 21.29 | 38.32 | 0.9273 | 0.9248 | 0.9340 | 0.9258 | 0.9340 | 0.9820 |
| ResNet50 | 23.51 | 42.82 | 0.9322 | 0.9302 | 0.9375 | 0.9307 | 0.9375 | 0.9816 |
| VGG16 | 134.27 | 164.19 | 0.9390 | 0.9365 | 0.9410 | 0.9371 | 0.9410 | 0.9839 |
| VGG19 | 139.58 | 207.68 | 0.8537 | 0.8111 | 0.8735 | 0.8295 | 0.8735 | 0.9725 |
| EfficientNet-B0 | 4.01 | 4.10 | 0.9278 | 0.9231 | 0.9323 | 0.9259 | 0.9323 | 0.9769 |
| MobileNetV3 | 4.20 | 2.35 | 0.9133 | 0.9090 | 0.9203 | 0.9115 | 0.9203 | 0.9745 |
| **Proposed method** | 11.36 | 18.98 | **0.9535** | **0.9495** | **0.9558** | **0.9521** | **0.9558** | **0.9854** |

The relatively low parameter count suggests that the proposed method can achieve its predictive performance without substantially increasing model size, while the reduced FLOPs indicate lower computational demands during the detection process. This computational efficiency is particularly advantageous for potential deployment in resource-constrained clinical environments, where computational cost and scalability may be important considerations. Overall, the results demonstrate that the proposed method provides an efficient alternative to more computationally intensive methods while retaining competitive predictive performance, potentially applicable to health-related mobile devices.

#### 4.2.6. External Validation

To further evaluate the generalizability and robustness of the proposed method, we performed external validation using an independent dataset collected from local clinics, which was not used during model development. The external cohort included 20 patients with AMD and 20 age- and sex-matched healthy controls. The same trained model was applied to the local dataset without additional model training or parameter tuning.

As summarized in Figure 9, the proposed method achieved more than 0.935 on all metrics, with a precision over 0.94 and an AUC over 0.99 at the image level. Similarly, the proposed method attained outstanding results at the patient level, as shown in Figure 10. Compared with the performance observed on the internal dataset, the model showed comparable performance on the external cohort.

**Figure 9.**
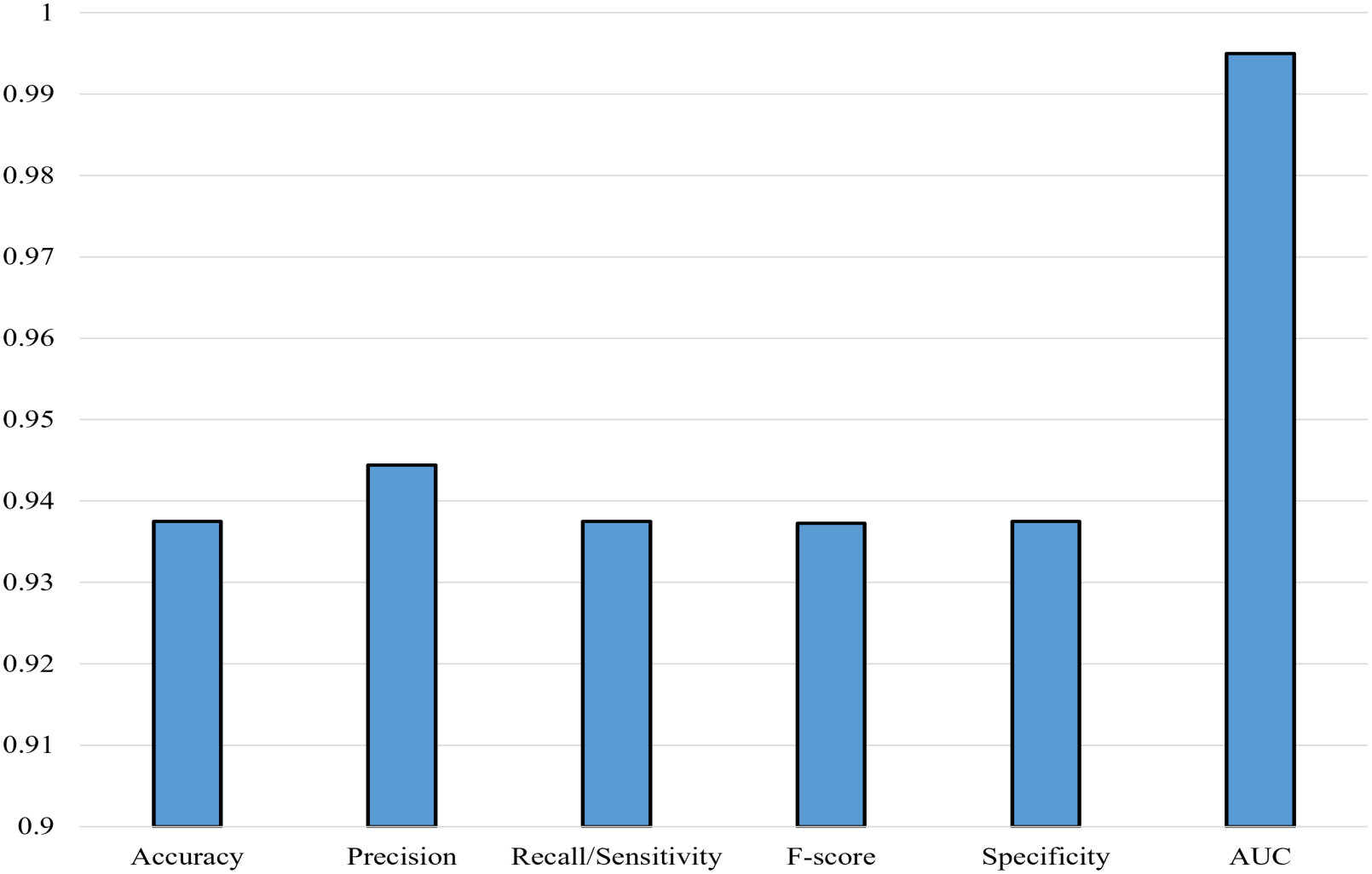
Image-level performance of the proposed model on the external cohort from local clinics

**Figure 10.**
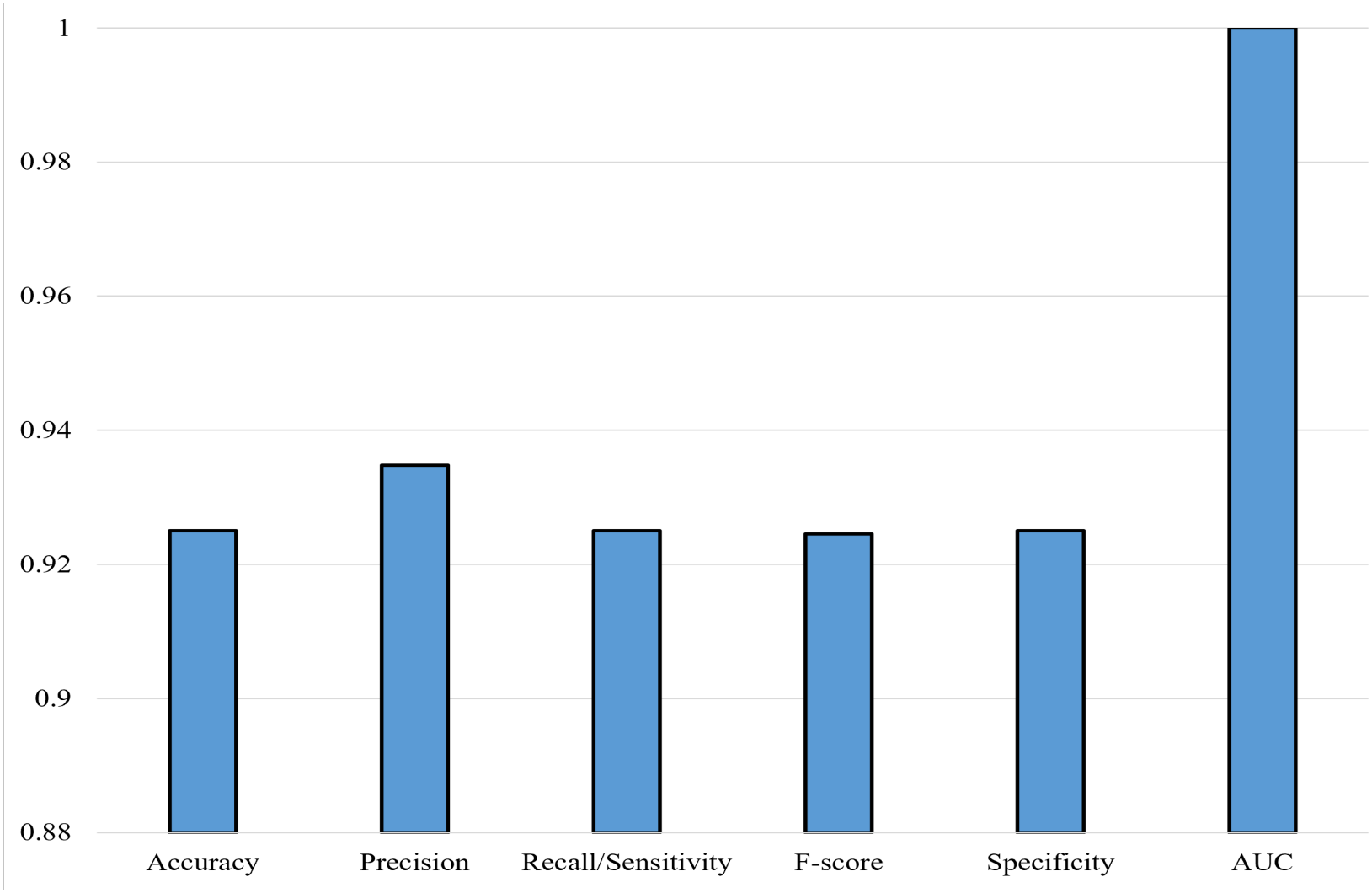
Patient-level performance of the proposed model on the external cohort from local clinics

The preservation of comparable performance on an independent dataset demonstrates that the proposed method does not solely depend on specific characteristics of the data used for model development. In particular, the model maintained high AUC values on the external cohort, suggesting that the learned representations captured clinically relevant patterns that remained informative beyond the original development dataset.

These findings provide evidence that the proposed method can generalize beyond the development dataset and maintain reliable performance under an independent data. The remaining performance gap may be attributable to differences in patient characteristics or data acquisition protocols, highlighting the importance of continued validation and evaluation across more heterogeneous clinical populations.

#### 4.2.7. Explainable AI

Explainable AI techniques are crucial for understanding the decisions made by the AI model. With the help of explainable AI, clinicians and patients are able to understand the prediction results of the proposed model more easily, thus fostering their trust in the AI model.

Gradient-weighted class activation mapping (Grad-CAM) [46] is used to generate heatmaps, highlighting important regions for disease prediction. The heatmaps are overlaid on the original images to visualize where the model focuses.

The explainable results of the correctly predicted examples by the proposed model are shown in Figure 11, where warm colors (such as red and yellow) denote higher importance, while cool colors (such as blue) indicate lower importance for disease detection.

**Figure 11.**
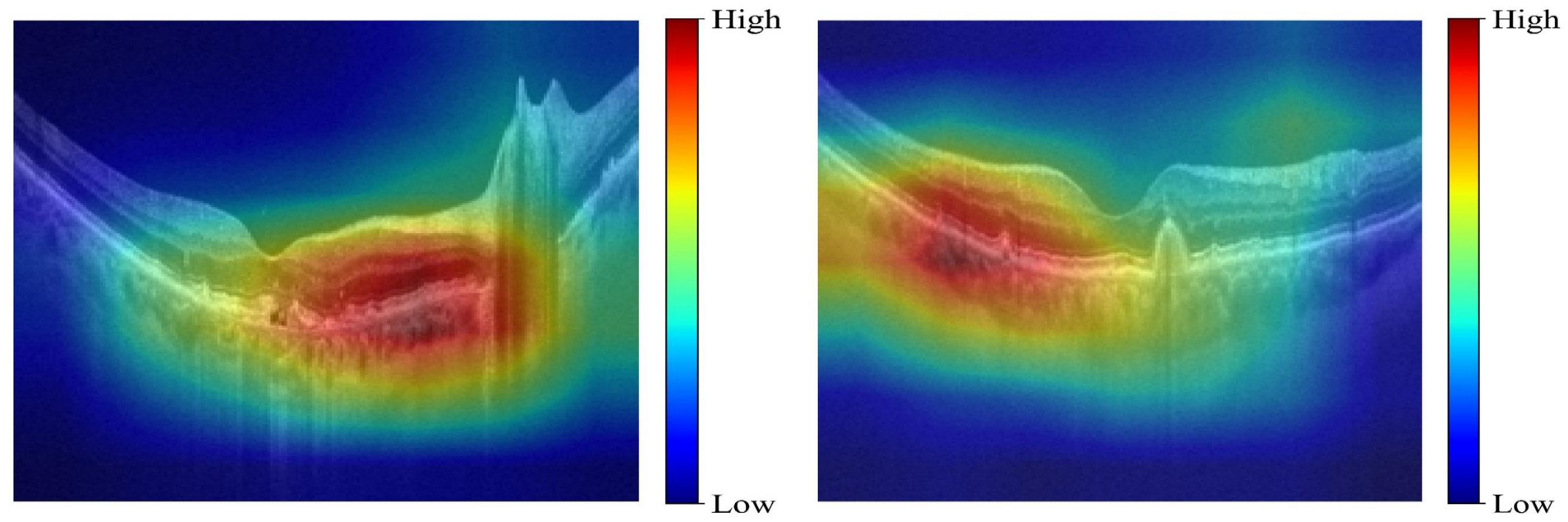
The correctly predicted examples by the proposed model

As illustrated in Figure 11, the proposed model has the ability to focus on the retinal layers within OCT images, which are the important regions for prediction. Clearly, the explainable results with heatmaps enable the proposed model to be clearer and more user-friendly, especially for patients and medical experts.

We were able to observe from experimental results that at the patient level, the model performance was comparable to that at the image level, suggesting that aggregating image-level predictions was still effective for disease prediction. The explainable analysis indicated that the proposed model had capability to focus on significant regions, such as retinal layers within OCT images. Model efficiency and generalizability analysis further demonstrated the feasibility of the proposed deep multimodal integration model. According to the experimental results, we found that the proposed model was capable of learning meaningful representations, illustrating that the use of the multimodal strategy is effective for disease prediction.

### 4.3. Clinical Relevance and Limitations

The clinical relevance of the proposed model lies in its ability to integrate information extracted from images with routinely available clinical metadata for target-class detection. Rather than replacing expert assessment, the model could potentially serve as an auxiliary decision support tool for disease detection, particularly in scenarios requiring the efficient screening of vast numbers of images. Notably, the use of patient-level predictions provides a more clinically relevant evaluation unit than image-level classification alone.

Despite its promising ability in detecting AMD, this study has several limitations. First, the size of the external validation cohort was relatively limited, which may lead to instability in performance evaluation. Consequently, the external cohort offers only a preliminary assessment of the model generalizability rather than definitive evidence of its clinical robustness. Future research should evaluate the model using larger, multicenter cohorts that encompass more diverse patient populations and data acquisition conditions across various hospitals and clinical institutions. The present evaluation remains unclear whether the model predictions can improve clinical decision-making or workflow efficiency. Prior to clinical implementation, further studies such as prospective multicenter validation, calibration assessment, clinical utility analysis, and workflow studies involving clinicians will be required. Such studies will help determine whether the model offers substantial benefits beyond existing clinical assessment.

In addition, it is noteworthy that the proposed AMD prediction model has great potential and applicability to help detect Alzheimer’s disease (AD) and Parkinson’s disease (PD), considering the availability of AMD OCT datasets and the strong association between AMD and AD/PD. To the best of our knowledge, currently available labeled OCT image datasets for AD or PD are extremely limited, making it highly challenging to directly train a reliable model for predicting AD or PD. Previous studies have discovered that AMD is clinically related to AD and PD, with AMD patients having an enhanced risk of developing AD and PD [47], [48], [49]. Specifically, AMD and AD/PD share some overlapping features, such as the thinning of the retinal layers, including retinal nerve fiber layer (RNFL) and ganglion cell inner plexiform layer (GCIPL). Image feature representations learned by AMD detection models, such as edges, layer contrast and patterns of thinning, can also be used for other tasks. These learned features can be transferred to the prediction of AD and PD via fine-tuning. Moreover, public OCT image data with annotated labels has been available for AMD. Hence, the development of the deep learning model for detecting AMD offers a great possibility to help predict AD and PD.

## 5. Conclusion

In this study, a deep multimodal integration model was developed to combine OCT images with clinical text data for AMD prediction. From extensive experimental results, the proposed model achieved superior performance both at the image level and at the patient level, with good accuracy, precision, recall or sensitivity, F-score, specificity, and AUC values. Visualization results based on ROC curves and confusion matrices, as well as heatmaps with an explainable AI technique, further showed the ability of the model to detect diseases. The proposed method demonstrated a good balance between predictive results and computational efficiency, with competitive performance and computational cost. Moreover, external validation on an independent dataset showed that the method maintained strong predictive performance beyond the model development dataset, supporting its robustness and generalizability. These findings highlight the potential of the proposed method as an efficient and generalizable solution for AMD detection. Based on a series of ablation analyses and model comparisons, our proposed method achieved excellent results in AMD detection compared with some existing methods by taking advantaging of a multimodal strategy.

In the future, the model would be further improved for better feature learning for disease prediction. One research direction is to fuse more modalities into the current model to obtain a deeper understanding of disease prediction. Despite yielding overall superior results on the external cohort, some performance variation was observed between the internal and external datasets, which may reflect differences in patient characteristics, data quality, or clinical practice. These findings highlight both the generalizability of the proposed method and the need for further validation in larger and more diverse patient populations.

### CRediT authorship contribution statement

**Ming Fang:** Conceptualization, Methodology, Validation, Formal analysis, Investigation, Data curation, Writing – original draft preparation, Writing – review & editing, Visualization. **Hamoon Miri:** Resources, Writing – review & editing. **Kerry Bishop:** Resources, Writing – review & editing. **Dorsa Robatjazy:** Data curation, Writing – review & editing. **Raymond C. Ko:** Resources, Writing – review & editing. **Ravikrishna Nrusimhadevara:** Resources, Writing – review & editing. **Francisco S. Cayabyab:** Writing – review & editing. **Seok-Bum Ko:** Conceptualization, Writing – review & editing. **Changiz Taghibiglou:** Conceptualization, Supervision, Funding acquisition, Writing – review & editing.

## Acknowledgements & Funding

The authors thank the Saskatchewan Health Research Foundation (SHRF) for providing research funding (#6595) to CT’s laboratory for this study.

## Data Availability

The OCTDL dataset is available at https://data.mendeley.com/datasets/sncdhf53xc/1.

The independent clinical dataset collected in this study is not publicly available due to patient privacy and confidentiality concerns.

## Ethics Approval

This study was approved by the University of Saskatchewan Biomedical Research Ethics Board (Bio-REB) (ID: 4760). Informed consent was obtained from all participants.

